# Disruption of Normal Saline Supply Chain due to Natural Disaster: an analysis of the impact of normal saline shortage on anesthesia practice in a large hospital system and models towards resiliency

**DOI:** 10.1101/2024.10.03.24312030

**Authors:** Rashid Hussain, Elizabeth Pickle, Taylor Lonjin, Janine Then, Ryan Rivosecchi, Jessica Cassavaugh, Mark Fichman, Ata Murat Kaynar

**Affiliations:** Virginia Commonwealth University, Division of Critical Care Medicine, Department of Anesthesiology, Richmond, Virginia, United States of America; University of Pittsburgh Medical Center, Department of Anesthesiology, Pittsburgh, Pennsylvania, United States of America; University of Pittsburgh Medical Center, School of Pharmacy, Pittsburgh, Pennsylvania, United States of America; Beth Israel Deaconess Medical Center, Department of Anesthesiology, Harvard Medical School, Boston, Massachusetts, United States of America; Carnegie Mellon University, Tepper School of Business, Pittsburgh, Pennsylvania, United States of America; University of Pittsburgh Medical Center, Department of Critical Care Medicine, Pittsburgh, Pennsylvania, United States of America

**Keywords:** Natural Disaster, Supply Chain, Normal Saline, COVID-19, Resource Allocation

## Abstract

In the wake of the COVID-19 pandemic and upheaval in the global supply chain, the healthcare sector has grappled with acute shortages of essential resources. Such shortages, while intensified recently in scale and frequency, are not new, as natural disasters have posed recurrent challenges. An illustrative example is the impact of Hurricane Maria in September 2017, which severely disrupted the production of normal saline (% 0.9 NaCl fluid bags) in Puerto Rico. Hospitals relying on “just in time” delivery models found themselves in a precarious situation, prompting a need for innovative solutions to sustain care delivery amidst the crisis. The occurrence underscores the vulnerability of healthcare infrastructure to external disruptions and emphasizes the need for adaptive strategies to ensure the resilience of the system in the face of unforeseen challenges.

Consequently, our focus shifted to an examination of the impact of Hurricane Maria on saline supplies at the University of Pittsburgh Medical Center (UPMC) towards building a resilient model. This study aims to elucidate how the flagship hospitals of the system navigated the challenges posed by the shortage, both at the individual provider level and on a broader, system-wide scale.

This investigation spanned an 18-month study period, encompassing the analysis of intravenous (IV) fluid demand and usage patterns before, during, and after the hurricane. Employing a multi-faceted approach, we integrated quantitative data with qualitative insights derived from three months of “participant observation” and survey data. This mixed-methods approach enabled us to see how individuals and systems within UPMC adapted to the crisis. Systems level adaptation occurred at UPMC’s operating room (OR), pharmacy and distribution supply chain levels, including by decreasing OR demand, while at the individual level healthcare providers changed their behavior by turning to alternative medications and carrier fluids. These adaptive measures undertaken at UPMC offer insights for future crises at both the organizational and individual levels within the healthcare system.

## Introduction

The pharmaceutical supply chain’s reliance on a “just in time” delivery model poses a vulnerability to hospitals with limited storage capacity for essential resources, particularly crystalloid solutions. This vulnerability became evident when Hurricane Maria struck Puerto Rico in September 2017, leading hospital systems, including the University of Pittsburgh Medical Center (UPMC), into a supply crisis.

In the Operating Rooms (ORs) of the UPMC Health System, where anesthesiologists and nurse anesthetists depend on readily available IV fluids for safe patient care, the shortage of 0.9% sodium chloride (normal saline or N.S.), posed a significant challenge. Our investigation focused on analyzing the demand and usage patterns of IV fluid before, during, and immediately after Hurricane Maria, shedding light on how our local healthcare system responded to surgical needs amid a disrupted supply chain.

Our findings are contextualized within the broader landscape of the U.S. healthcare system, which grapples with persistent drug shortages. The Food and Drug Administration (FDA) defines drug shortages as instances where the demand for a drug exceeds its supply within the U.S. Sterile injectables and parenteral medications are particularly susceptible, given their high production costs and the stringent requirements for maintaining manufacturing facilities. Notably, compliance issues with regulatory standards at manufacturing facilities have contributed to significant shortages in the past.

Since the onset of the COVID-19 pandemic, these supply strains had become even more pronounced, with heightened demand for critical supplies like alcohol hand sanitizers, face masks, and intermittent shortages of various other medical supplies. In the aftermath, the healthcare system has continued to be plagued by shortages. This underscores the urgency of learning from case studies such as ours, as they offer insights into adaptive strategies and innovations that can enhance the resilience of healthcare systems in the face of acute demands and supply chain disruptions.

## Materials and Methods

The UPMC Q.I. committee approved the project entitled “The pattern of use of IV fluids in the operating rooms before and after nationwide IV fluid shortage and associated complications - Project ID: 1445.” We collected the data needed to study the crisis response from the two major teaching hospitals of UPMC-- Presbyterian (PUH) and Montefiore University Hospital (MUH) hospitals, which have a total capacity of 750 medical and surgical beds, 150 ICU beds, and 40 anesthetizing locations including ORs, cystoscopy and bronchoscopy suites.

Data collection encompassed OR usage of crystalloid from central databases and qualitative survey responses from operating room providers in the Department of Anesthesiology and Perioperative Medicine. We chose the study period of 18 months from April 2017 to the start of October 2018 to include IV fluid demand and usage before, during, and after Hurricane Maria in September 2017. We collected IV fluid “demand” data (requested volume and supply by OR pharmacy) for both small (≤500 mL) and large volume (>500 mL) IV fluid from the supply management and pharmacy services at UPMC for the 18-month study period. Data on “usage” were collected from the electronic medical records, pharmacy charges, and supply chain management. To elucidate how the UPMC system responded during this shortage, we conducted surveys and analyzed using qualitative methods. We did survey anesthesia providers involved in the administration of IV fluids and our questions focused on the use of IV fluids in the OR, use patterns in the OR, what they were used for, alternative solutions, and perceptions of how the shortage impacted care.

## Results

### Operating Room IV Fluid Demand and Usage Data

The demand data for small and large IV fluid bags over the 18-month study period are presented in Figures 1 and 2. The hurricane started on September 16^th^, 2017, and major storm activity ceased on October 2^nd^, 2017. Small volume IV fluid bag demand began to decrease in October 2017. Strain in the supply was detected by the UPMC supply management system and was readily apparent in November when the full-scale implementation of alternatives had taken place. The total number of small-volume IV fluid bags supplying PUH & MUH operating rooms decreased dramatically from 1,652 in October 2017 to zero in November 2017. It remained depressed well below pre-hurricane numbers until supply began to increase again in June 2018.

**Figure 1a:**
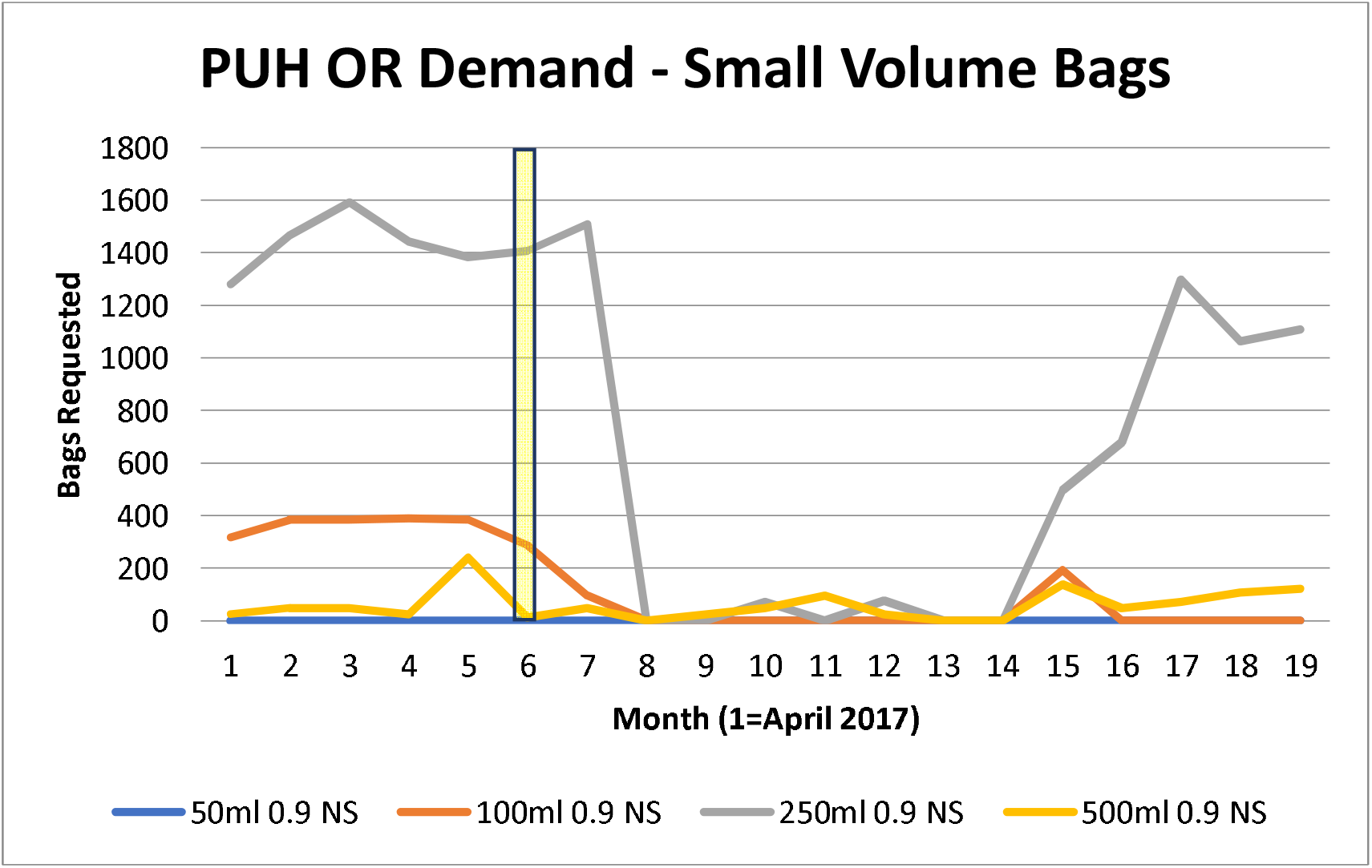
The impact of small volume (≤500 mL) IV fluid use with the Hurricane Maria at Presbyterian Hospital. Month of Hurricane Maria Highlighted.

**Figure 1b:**
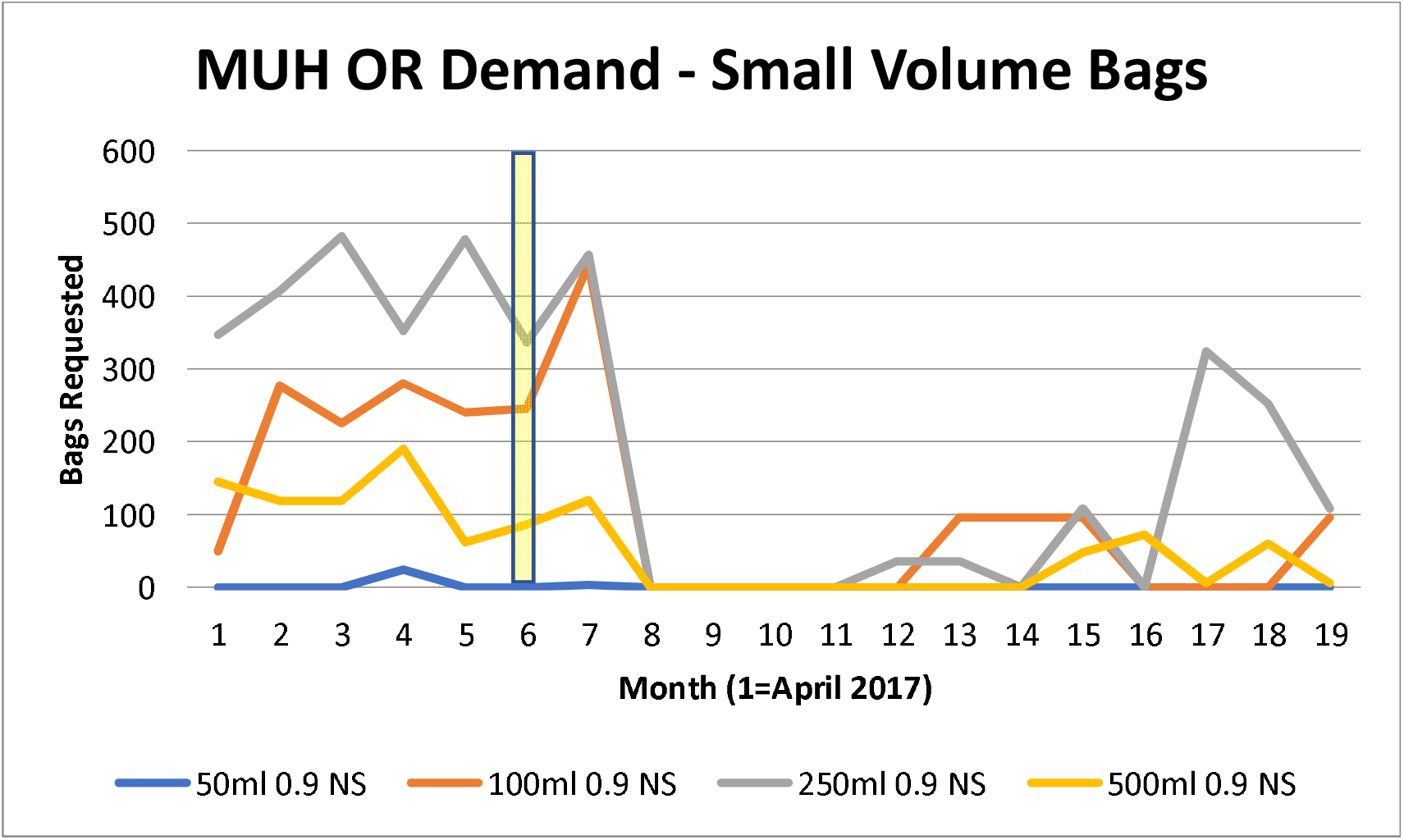
The impact of small volume (≤500 mL) IV fluid use with the Hurricane Maria at Montefiore Hospital. Month of Hurricane Maria Highlighted.

**Figure 2a:**
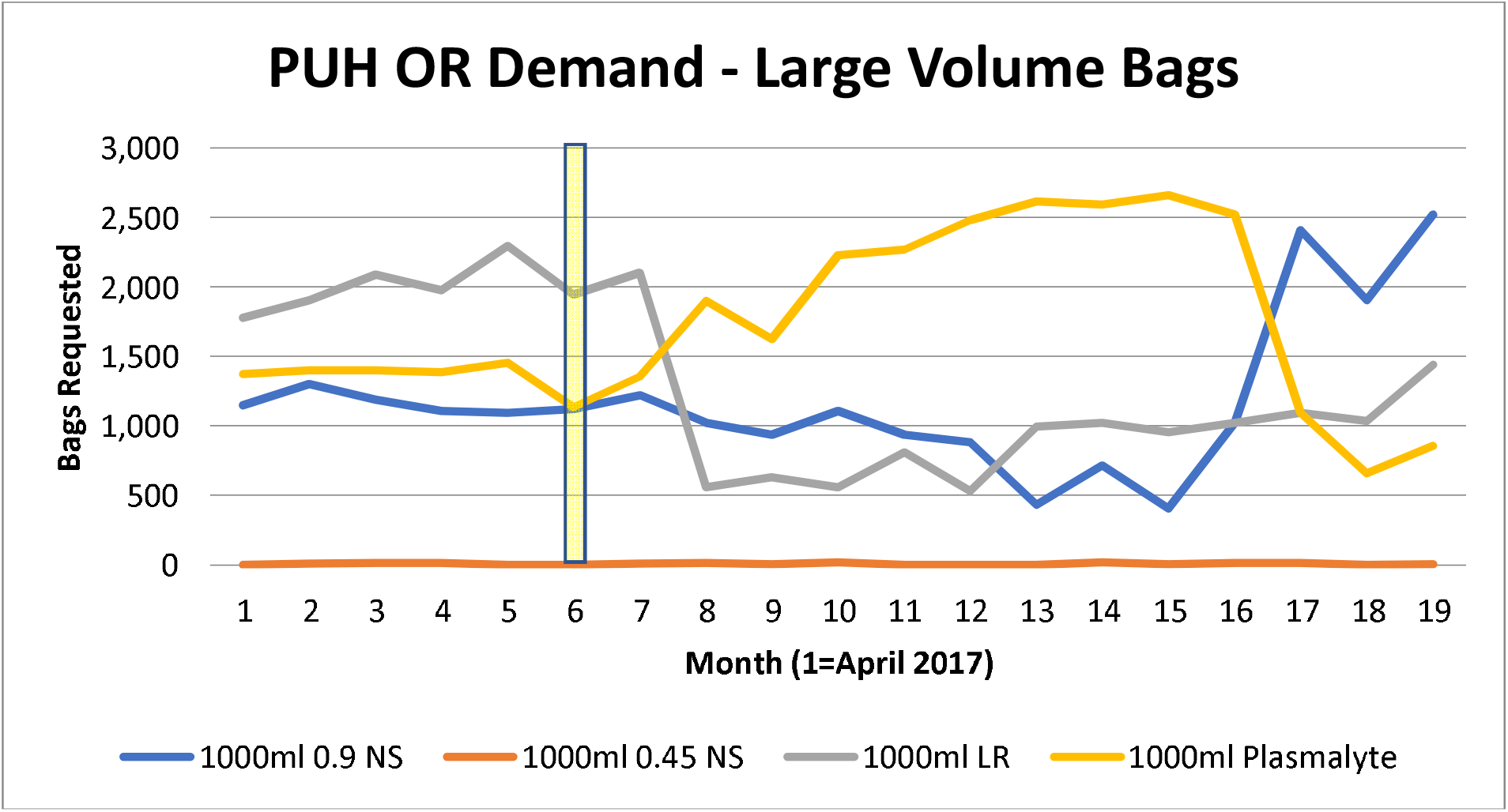
The impact of large volume (1,000 mL) IV fluid use with the Hurricane Maria at Presbyterian Hospital. Month of Hurricane Maria Highlighted.

**Figure 2b:**
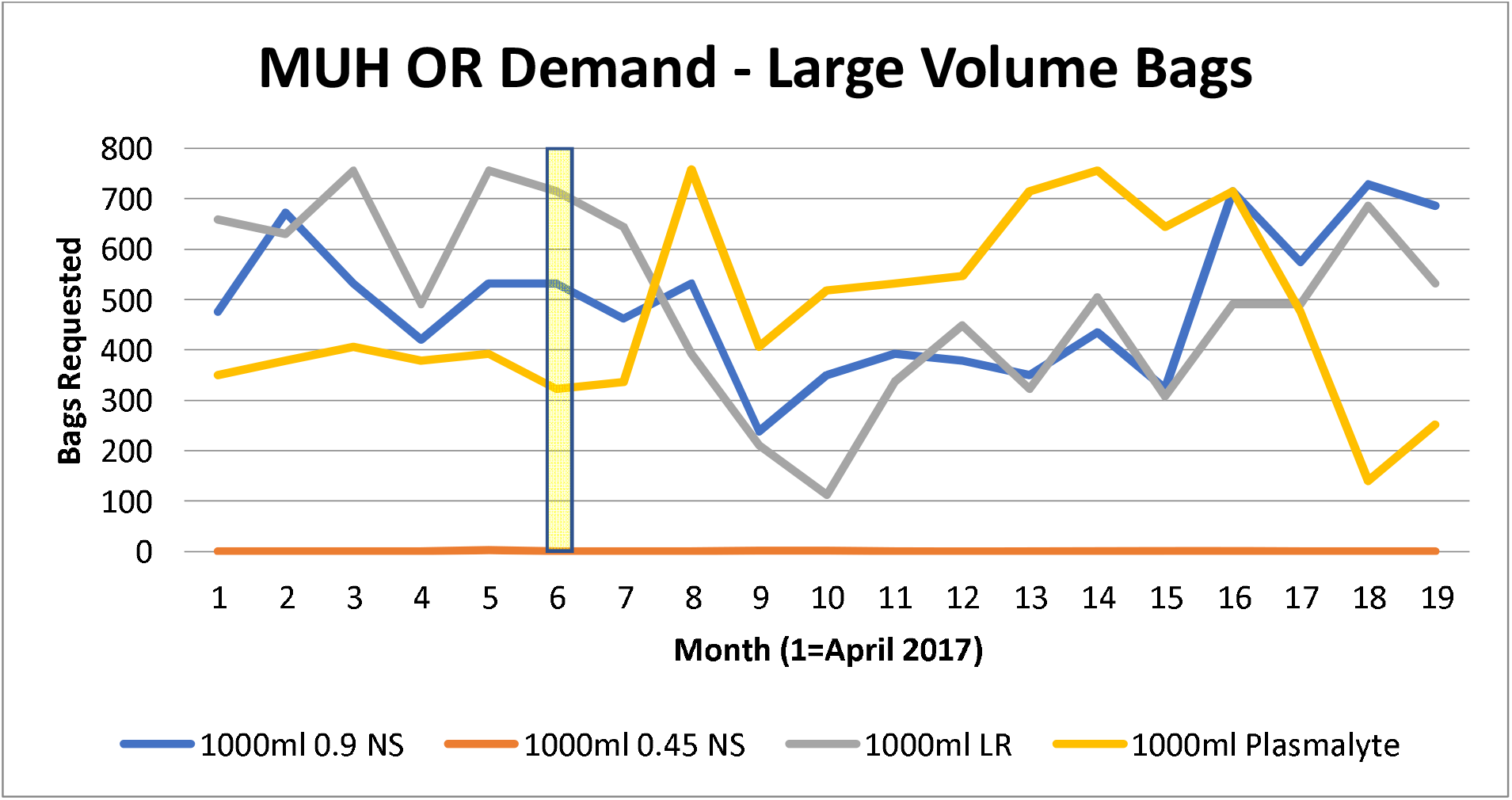
The impact of large volume (1,000 mL) IV fluid use with the Hurricane Maria at Montefiore Hospital. Month of Hurricane Maria Highlighted.

At PUH, this trend appeared to be primarily driven by changes in 100 mL and 250 mL bags. For example, the 100 mL bags ranged from 388 bags/month in July 2017 to 288 bags/month in September 2017, 96 bags/month in October 2017, and remained zero after that until June 2018 (Figure 1a). Small volume IV fluid bags followed a similar trend at MUH, with all small volume bag types diminishing to zero in November 2017 until supply began to increase in March 2018 (Figure 1b) slowly. The total demand for large volume IV fluids remained relatively unchanged through the 18 months at both PUH and MUH; however, the choice for IV Plasmalyte 148, also known as Plasma-Lyte A, did almost double at both locations, with an inflection point in January 2018 at PUH and in November 2017 at MUH (Figures 2a and 2b, respectively). Demand for large volume Lactated Ringer’s (L.R.) solution appeared to experience a mild decline starting in November 2017 at both hospitals. Large-volume normal saline demand remained relatively stable at MUH until it decreased in December 2017. Demand for large volume 0.45% sodium chloride or half-normal saline, also declined at both PUH and MUH during the study period, although less severely than small volume bags with less usage at baseline (Figure 2).

### Qualitative and Survey Data

A total of 80 unique anesthesia providers (nurse anesthetists and anesthesiologists) who were self-selected to have been present during the shortage responded to the survey from the total anesthesiologist, resident, and nurse anesthetist staff at UPMC Presbyterian and Montefiore. The survey comprised of questions on utilization of large and small normal saline IV fluid bags.

### Drug Shortage Survey Data

One question asked participants, “Which medications were on shortage that you felt most adversely affected your care?” Survey respondents reported that their experience of shortages was related to normal saline (21.25%, n=17), lidocaine (11.25%, n=9)—both specifically affected during Hurricane Maria—, narcotics (21.25%, n=17), sodium bicarbonate (3.75%, n=3), and “other” (41.25%, n=33), which included responses such as bupivacaine spinals, beta-blockers, vasopressin, and ketamine (drugs periodically on shortage generally).

Some of the key comments regarding the shortages (in general and specific to fluid) included:

- “We often have medication shortages that result in my using either non-optimal drugs or drugs that I am less familiar with to treat our patients.”
- “Not having availability to the best medication options for our patients. Not having IVF bags and treating a lot of hypotension.”
- “I have had to choose medications that were not ideal for my patients’ care.”
- “The decease (sic) or lack of remifentanil does alter the care of some of my neuro-anesthesia care. The saline shortage with not being able to hang saline on all G.I. patients so even today the G.I. lab no longer uses maintenance fluids for their cases not even colons who had preps…”

### Small Saline Bag Survey Data

Small volume saline bags are commonly used to dilute and deliver medications. In response to a question regarding their use of small volume saline bags, 60% of respondents reported using small saline bags daily. When asked for what purpose they used small saline bags, 13.75% (n=11) of respondents replied they used them for insulin, 18.75% (n=15) responded for antibiotics, 5% (n=4) for ketamine, and 50% (n=40) responded “other” with the comment field revealing that 45% of those specifically mentioned remifentanil, 10% responding with a vasopressor, and the remainder listing magnesium or dilute medications generally. A follow-up question asked what their alternative crystalloid of choice was, to which 16.25% (n=13) responded D5W, 12.5% (n=10) responded D5LR, and 51.25% (n=41) responded “other.” Under “other,” respondents did not have a uniform or trend in responses for alternatives, writing in a range of responses, from “none” to “pharmacy prepared bags” to “unsure” to “larger bags.”

### Large Saline Bag Survey Data

Though the large normal saline bag supplies were less strained, we collected IV fluid use responses and changes in their use patterns. Regarding the purpose of use, there was limited scope for the use of these larger bags: 10% (n=8) of respondents replying it was their “routine crystalloid,” 25% (n=20) responding it was used as a carrier with blood transfusions, 20% (n=16) responding it was continued from pre-op, 5% (n=4) using it with hyperkalemic patients, 2.5% (n=2) using it to help start an IV, and 33.75% (n=27) responding “other” --with 15% of those reporting use for a neurosurgical case and the rest declining to use it at all. When asked about frequency of use of the larger saline bags, 36.25% (n=29) responded never using it at all, 15% (n=12) monthly, 31.25% (n=25) weekly, and 15% (n=12) daily. Additionally, for their alternative crystalloid, respondents overwhelmingly responded Plasma-Lyte at 53.75% (n=43), followed by lactated ringer solution (LR) at 31.25% (n=25) and 10% (n=8) choosing “other” writing “Plasmalyte or LR” in the comments section.

Specific medications in the OR affected (in terms of usage) included remifentanil, a rapidly esterized opioid used for neurosurgical cases and has to be mixed before a case usually, ketamine used for chronic pain patients, and an adjunct in Enhanced Recovery After Surgery (ERAS) protocols, insulin, and antibiotics. Surgical load as determined by hours of OR time decreased in a planned manner, correlated with the time of the hurricane (See Figure 3), providing some relief during the crisis. Nonetheless, innovation and adaption were implemented as seen in the survey at the level of bolus dosing medications used previously in infusions, providers making their own syringe-pumps for drugs, or relying on new pharmacy preparations which took planning to implement (ex: order the night before to ensure availability for scheduled case).

**Figure 3a:**
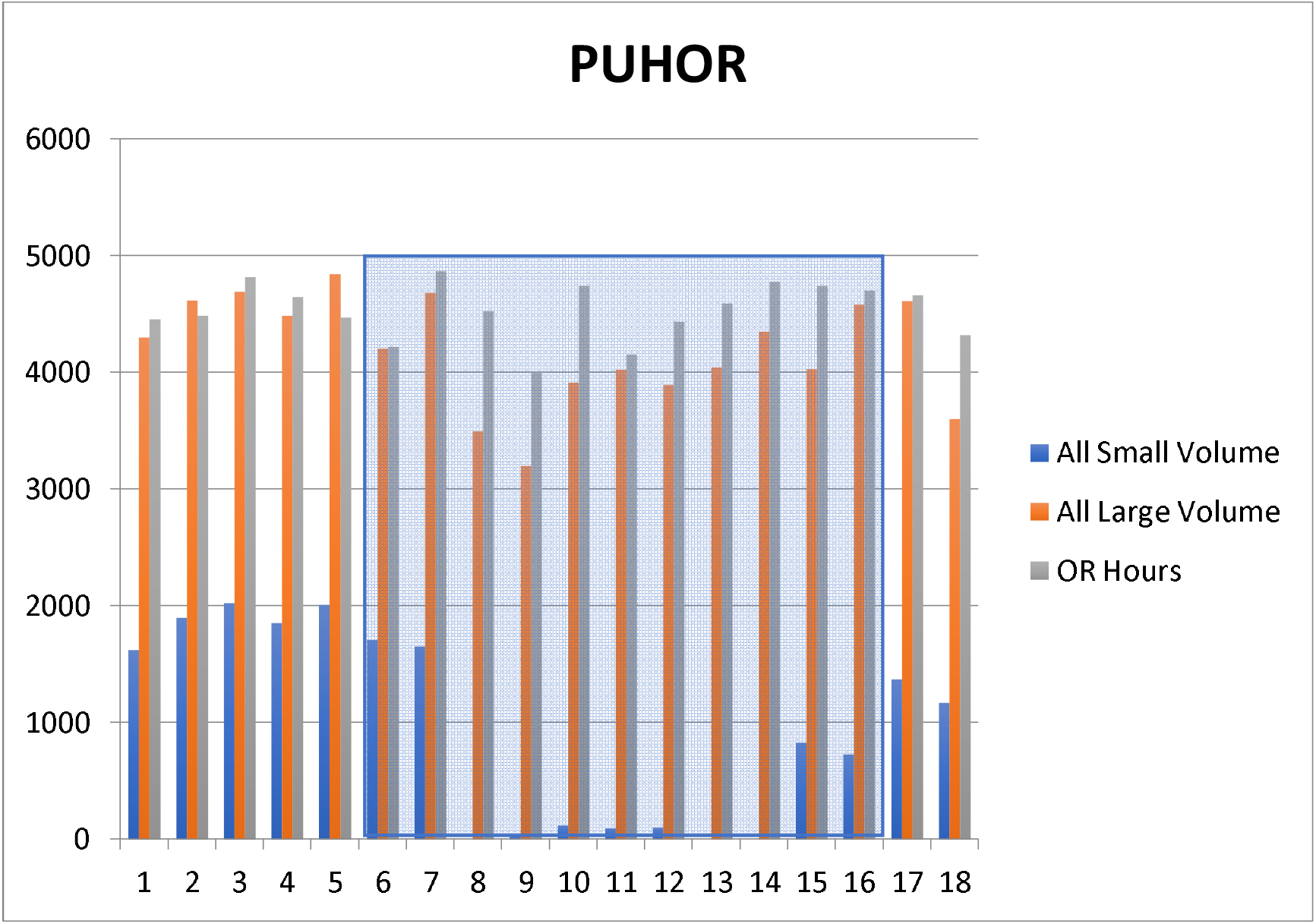
Operating Room Hours Logged Per Month at Presbyterian Operating Rooms (O.R.s) along with total number of Bags of Crystalloid (Small and Large). Shortage Months Highlighted.

**Figure 3b:**
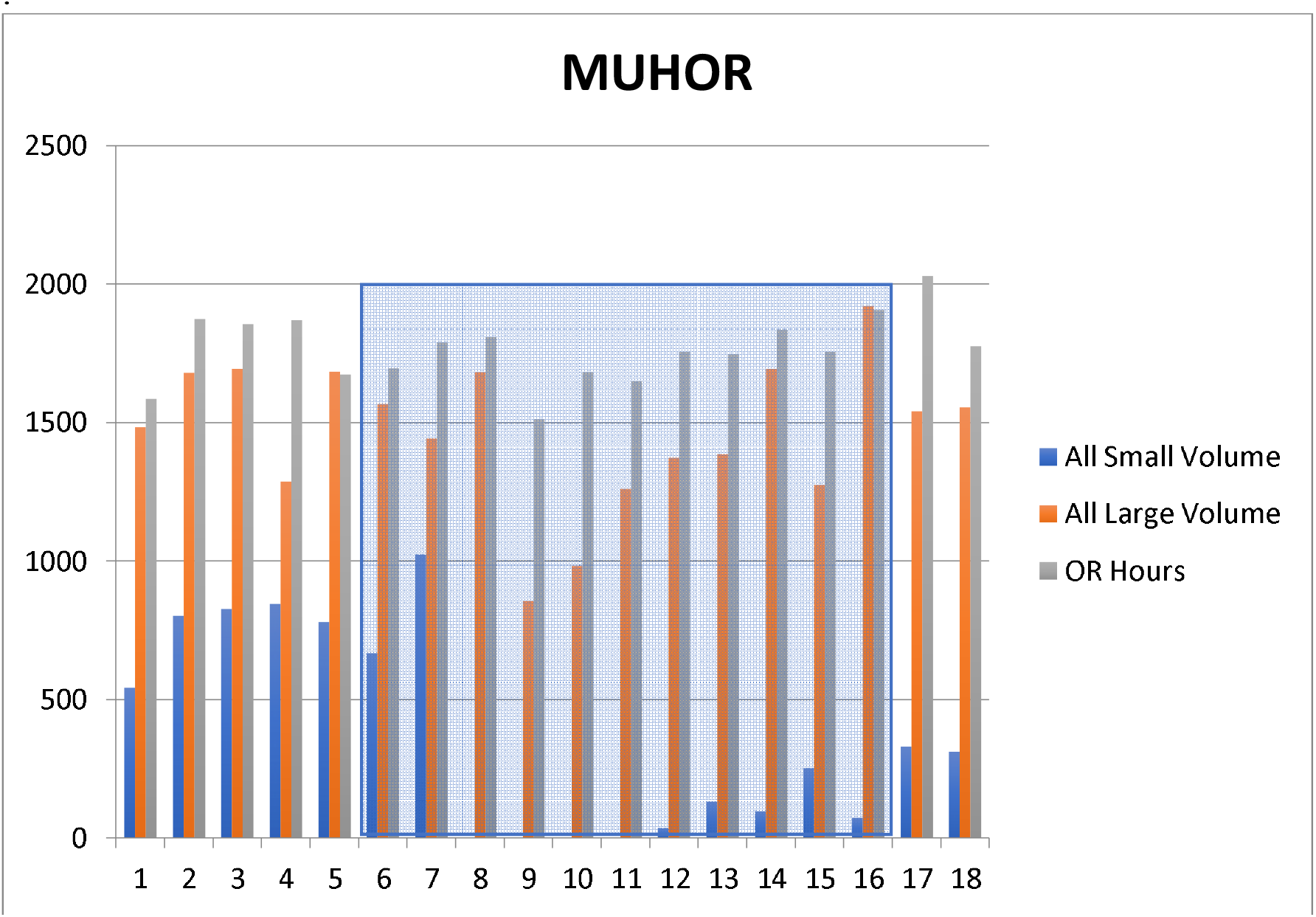
Operating Room Hours Logged Per Month at Montefiore Operating Rooms (O.R.s) along with total number of Bags of Crystalloid (Small and Large). Shortage Months Highlighted.

## Discussion

We present a case study detailing the healthcare system’s response to a critical shortage of small bags of normal saline in the aftermath of a natural disaster. The crisis had a dual origin, stemming from chronic shortages in the parenteral drug and fluid supply, exacerbated by the immediate impact of an environmental disaster—specifically, Hurricane Maria. The adverse effects were particularly pronounced in the U.S. supply of small bags, defined as 250 mL or less (with small volume parenteral being 100 mL or less) of normal saline.^6^ Baxter, a leading IV fluids manufacturer, temporarily ceased operations at all Puerto Rican plants for a month, with power restored only by November 2017.^7-8^ UPMC, operating normally in August 2017, found itself unexpectedly facing a disruption in saline bag supply due to the hurricane—an unforeseen and “random” event. This natural disaster provided a unique opportunity to analyze the repercussions of such seemingly arbitrary occurrences. Throughout this period, the strain on anesthesia supply services necessitated adaptive strategies until a sustainable solution could be implemented.

Adaptation during this crisis manifested at various levels, encompassing both systems-based adjustments within pharmacy and supply chain sectors, as well as individual responses, particularly notable among anesthesia providers in the operating room. The emphasis on adaptability in crisis situations can foster innovation to ensure the survival of the system, albeit with an inherent risk of potential errors.

Individual adaptation by anesthesia providers in the operating room revealed a heightened risk of errors, such as drug dilution and microbial contamination. The shift to individual mixing outside the controlled environment of a compounding pharmacy raised concerns. Survey participants, when queried about preferred alternatives to small volume saline bags for compounding medications, presented diverse responses, indicating a lack of uniformity in their coping strategies during the shortage crisis.

Notably, providers expressed challenges in safely administering anesthetics for neurosurgical cases, where the customary practice of mixing remifentanil infusions in small saline bags was disrupted by shortages. The reliance on small saline bags as a compounding source for crucial medications like antibiotics, ketamine, insulin, vasopressors, and remifentanil became evident. In response, providers resorted to mixing medications in alternatives or substituting with alternative medications, such as opting for long-acting narcotics and increased use of propofol in lieu of remifentanil. Interestingly, for ketamine and insulin, a systemic solution emerged with pharmacy pre-mixing bags, demonstrating a collaborative approach. The increased regulation of medications through pre-mixed or compounded bags from the central pharmacy mitigated challenges associated with compounding during the shortage, offering a systemic remedy to address individual concerns and enhance overall safety protocols.

Also at the individual level, a noticeable decline in demand for 1L normal saline bags was observed, despite not being officially declared in shortage. Instead, there was a discernible shift towards alternatives such as Lactated Ringer and Plasma-Lyte. This trend coincided with recent clinical trials, specifically the SMART-SURG and SMART-MED studies, focusing on the impact of crystalloids on surgical and medical outcomes.^9-10^ These trials, well-known during the study period, provided support for considering alternative crystalloids over normal saline.

The Isotonic Solutions and Major Adverse Renal Events Trial in Medicine (SMART-MED) and Surgical patients (SMART-SURG) investigated critically ill medical and surgical ICU patients, comparing the outcomes of normal saline versus balanced crystalloids. The primary endpoint was a major adverse kidney event within 30 days, defined as death, new renal-replacement therapy, or persistent renal dysfunction. The results showed a slight difference in major adverse kidney events (15.4% in normal saline vs. 14.3% in balanced crystalloids groups, p=0.04). Notably, acute kidney injury rates suggested that the use of balanced crystalloids could be equally effective in resuscitation. Some anesthesia providers acknowledged that these studies influenced their choice of crystalloid, reflecting a growing awareness and consideration of the evolving evidence base in their clinical decision-making processes.

Systemic solutions during this acute crisis have notably focused on two key areas: the operating room (OR) pharmacy’s provision of standardized solutions and the broader system supply chain at UPMC.

The emphasis on the OR pharmacy providing standardized solutions has proven crucial in addressing the challenges posed by the shortage. By preparing standardized solutions, the pharmacy significantly reduces the risk of dilution errors and contamination. This standardized approach not only enhances patient safety but also streamlines the process of adopting alternative solutions in a timely manner.

The OR pharmacy, in this context, has emerged as a strategic player in improving the delivery of critical medications and actively contributing to closed-loop planning for scheduled cases. The study’s findings highlight the positive impact of this approach, particularly with the standardization and preparation of Ketamine and Insulin by the local pharmacy.

Expanding on this concept, the broader idea of localizing or “reshoring” pharmaceuticals within the continental US mainland or even more specifically local hospital and healthcare system framework emerges as a proactive strategy. This approach aims to mitigate the impact of acute shortages by establishing a healthcare system-driven pharmaceuticals model, thereby increasing the availability of high-quality pharmaceuticals.

The significance of localization became evident during Hurricane Maria when Baxter’s facilities were incapacitated, leading to the depletion of national reserves on the U.S. mainland.^7^ Subsequently, months later, the situation improved with increased supply from alternative manufacturers like B. Braun Medical and ICU Medical, as well as the establishment of alternative production sites, including Brazil by January 2018.^3^

Consideration of pharmaceutical localization aligns with the vision outlined by U.S. Homeland Security, envisioning large hospital systems such as UPMC and Kaiser Health taking on the production of key medications or medical supplies. While this proactive measure could enhance the healthcare system’s resilience, startup costs and the ongoing maintenance of such facilities on the mainland have been primary arguments against pursuing this strategy.^11^

Other systems solution during this crisis included, for example, the decrease in demand. This was done with decreased case volume, as seen in Figure 3 with surgical load. Elective cases were decreased though reason for this being the hurricane itself is unknown due to the retroactive nature of aspects of this study. Notably though, all providers were made aware of the crisis and shortage affecting supply and need with the resultant drop reflecting a systemic change in planning.

### Lessons Learned

Unfortunately, the frequency of disasters is projected to rise in the coming decades due to global environmental changes.^12^ The recent pandemic of COVID-19 exemplified the strain on the supply chain for sedatives and analgesics,^13^ compelling hospitals and providers to rapidly explore alternative sources or even resort to in-house manufacturing, often under non-standard hygienic conditions.^4^ As discussed, while the idea of health systems producing their own drugs presents challenges, particularly in terms of upfront investment and costs, local production supported by government initiatives could offer cost savings and strategic advantages.^14^

Furthermore, this study underscores the crucial role of well-trained healthcare providers in building resilience into the system. Although providers can take initiative individually, implementing system controls with feedback, such as pharmaceutical oversight of compounding with streamlined processes, is essential for ensuring the safe and timely delivery of medications. The study also reveals that resource limitations prompted a shift towards safer balanced crystalloids, aligning with recommendations in the literature. These changes, initiated at the individual level, contribute to systemic adaptations, while shifts in case volume demonstrate a bottom-up influence on the system.

Importantly, in response to natural disaster challenges, the American Medical Association (AMA) and the United States Pharmacopeia (USP) recently published recommendations that hospital systems can take to overcome supple chain shortages. These include: 1) Incentivize advanced manufacturing technology and develop new continuous manufacturing technology for critical drugs and active pharmaceutical ingredients; 2) Improve the function and composition of the Strategic National Stockpile; and 3) Improve multinational cooperation on supply chain resilience.^15^

In future long-term planning, it is essential to integrate a systematic strategy that allows the healthcare system to adapt to individual-level changes. We present a framework, depicted in Figure 5, which outlines the steps to construct a resilient healthcare system. Based on our experience working with health systems and suppliers, leaders can embrace four pivotal initiatives to strengthen their supply chains. First and foremost, enhancing visibility within the supply chain is crucial.

**Figure 5.**
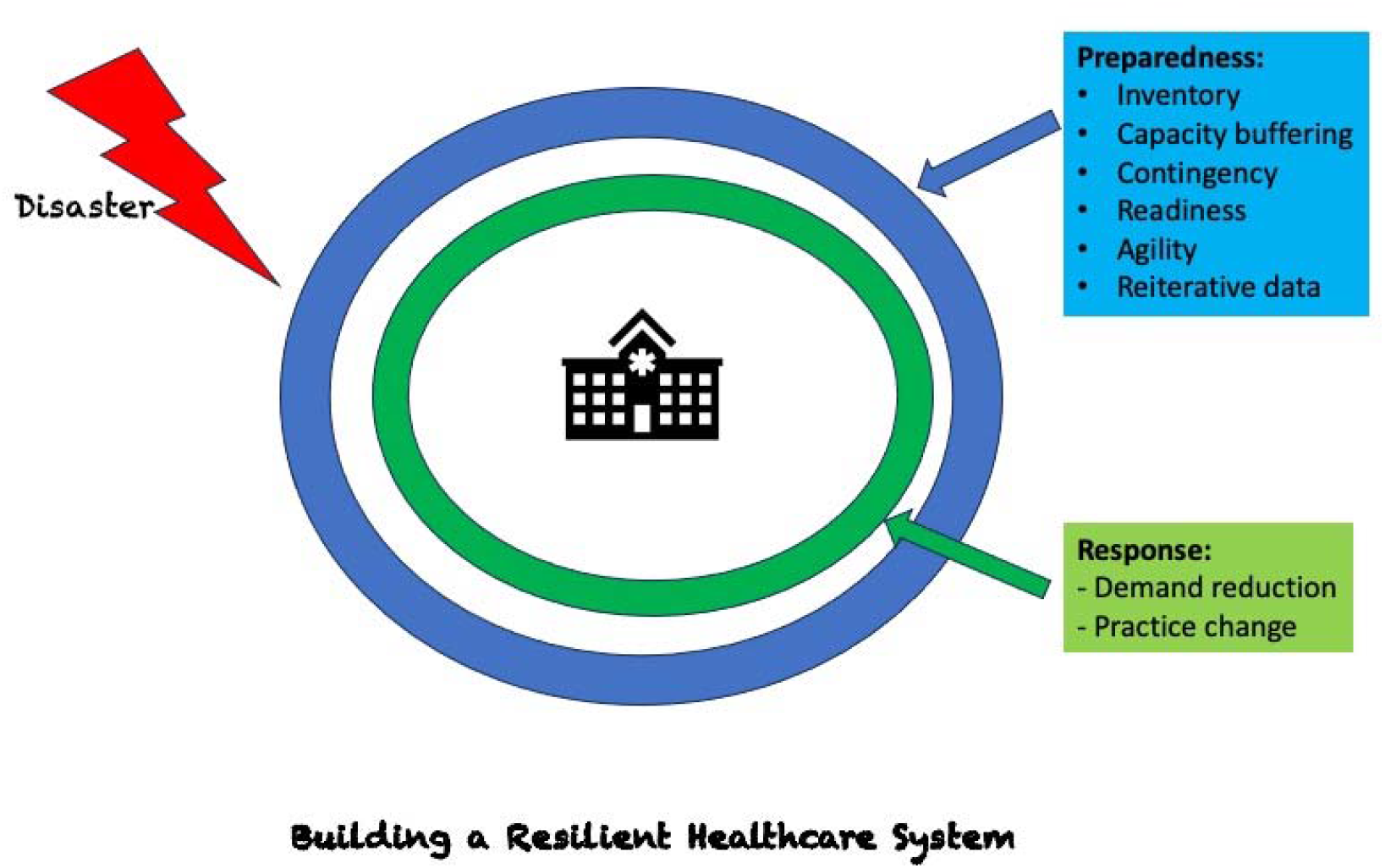
Illustrative concept for building a Resilient Healthcare System.

Internally, this means consolidating inventory data and utilizing innovative tools like RFID barcoding to improve comprehension of inventory levels across different care settings. Externally, partnering with group purchasing organizations (GPOs) and distributors provides valuable insights into supply chain dynamics, facilitating early identification of potential disruptions.^16^ Secondly, health systems should adopt product-specific strategies to address supply chain disruptions. It is vital to pinpoint critical items through collaboration with clinicians, emergency preparedness teams, GPOs, and distributors.^16^ Subsequently, proactive actions can be initiated, such as establishing demand management protocols and exploring alternative products. Moreover, considering stockpiling strategies for essential items can mitigate the risk of shortages and serve as a protective measure during crises. Lastly, the development of pertinent protocols, capabilities, and governance structures is indispensable for efficient supply chain management.^16^ Health systems need to set clear guidelines for product utilization in partnership with stakeholders and allocate resources to respond promptly to disruptions, including adjusting demand and adapting practices. Strong governance mechanisms ensure efficient decision-making, which is crucial for agile responses to supply chain issues. By optimizing expenditures and maintaining a lean financial base, health systems can reinforce their resilience. This approach allows them to withstand potential challenges and ensure the continuous provision of vital healthcare services without interruption.

### Future Directions

The scope of this paper was limited to two university hospitals in a multihospital system. Further investigation encompassing multiple hospitals, or the entirety of the health system would provide deeper insight into healthcare systems’ capacity to cope with drug shortages. Healthcare systems must operate under the assumption that future events like these will occur. Reports such as our investigation highlight the importance of a multi-pronged approach to innovation and solutions in the face of shortages so that healthcare entities from a pharmacy, supply management, or providers can creatively approach these issues. Further research and efforts are needed to address these inevitable crises.

### Limitations

This paper provides qualitative descriptors and OR demand and usage data to present how a health system responded to shortages generally and a severe shortage more acutely. The surveys (2019) were completed two years after the hurricane (2017). The survey data would thus be vulnerable to recollection bias and selection bias by design, though not directly observed. The study was not validated externally to other healthcare systems. Additionally, the data is limited in its specific scope of supply, demand, and usage and cannot account for different causes, some of which were described in the discussion.

## Data Availability

All data produced in the present work are contained in the manuscript

## References

1. Alphonse J, Bellam S, Fernandez M et al., The FDA funding crisis. J Pharm Technol. 2014; 30:57□60.

2. Tucker EL, Cao Y, Fox ER, and Sweet B. The drug shortage era: a scoping review of the literature 2001–2019. Clinical Pharmacology & Therapeutics. 2020; 108:1150–1155.

3. Mazer-Amirshahi M, Fox ER. Saline shortages - many causes, no simple solution. N Engl J Med. 2018; 378:1472□1474.

4. Peters G. U.S. Senate Committee on Homeland Security & Governmental Affairs (HSGAC). A price too high: cost, supply, and security threats to affordable prescription drugs (December 2019). https://www.hsgac.senate.gov/download/191206_report_apricetoohigh (Accessed 2021 May 19).

5. Pasch RJ, Penny AB, Berg R. National hurricane center. Tropical cyclone report: hurricane Maria (April 2018). https://www.nhc.noaa.gov/data/tcr/AL152017_Maria.pdf (Accessed 2021 May 19).

6. American Society of Health-System Pharmacists and University of Utah Drug Information Service. Small-volume parenteral solutions shortages: suggestions for management and conservation (October 2017). https://www.fda.gov/media/108408/download (Accessed May 2021).

7. Sacks CA, Kesselheim AS, Fralick, M. The shortage of normal saline in the wake of hurricane Maria. JAMA Internal Medicine. 2018; 178: 885.

8. Koons C, Langreth R. Facing saline shortage, FDA prioritizes some Puerto Rico plants. Bloomberg – Industry Week (Nov 2017). https://www.industryweek.com/leadership/article/22024587/facing-saline-shortage-fda-prioritizes-some-puerto-rico-plants (Accessed 2021 May 19).

9. Semler MW, Self WH, Wanderer JP et al., Balanced crystalloids versus saline in critically ill adults. N Engl J Med. 2018; 378:829□839.

10. Burdett E, Dushianthan A, Bennett-Guerrero E et al., Perioperative buffered versus non-buffered fluid administration for surgery in adults. Cochrane Database Syst Rev. 2012; 12:CD004089

11. Abelson R, Thomas K. Fed up with drug companies, hospitals decide to start their own. New York Times. January 18, 2018. https://www.nytimes.com/2018/01/18/health/drug-prices-hospitals.html.

12. Ting M, Kossin JP, Camargo SJ et al. Past and future hurricane intensity change along the U.S. east coast. Sci Rep. 2019; 9:7795.

13. Ammar MA, Sacha GL, Welch SC et al., Sedation, analgesia, and paralysis in COVID-19 patients in the setting of drug shortages. J Intensive Care Med. 2021; 36:157–174.

14. Gurvich VJ, Hussain AS. In and beyond COVID-19: U.S. academic pharmaceutical science and engineering community must engage to meet critical national needs. AAPS PharmSciTech. 2020; 21:153.

15. Anesthesia Incident Reporting System Case 2024-02: Supply and Demand – Importance of the Supply Chain. ASA Monitor 2024; 88:1–11

16. National Academies of Sciences, Engineering, and Medicine; Health and Medicine Division; Board on Health Sciences Policy; Committee on Security of America’s Medical Product Supply Chain; Shore C, Brown L, Hopp WJ, editors. Building Resilience into the Nation’s Medical Product Supply Chains. Washington (DC): National Academies Press (US); 2022 Mar 3

